# Compound heterozygous DAW1 variants reveal tissue-specific roles in left-right patterning and congenital heart disease without primary ciliary dyskinesia

**DOI:** 10.64898/2026.01.27.26344789

**Authors:** Dana Urbatsch, Anburaj Jeyaraj, Shruti Bedekar, Venkatramanan Rao, Shelby C. White, Matthew J Thomas, Andrea Garrod, Christina Peroutka, Aakrosh Ratan, Saurabh S. Kulkarni

**Author notes:** Lead author Saurabh S. Kulkarni, University of Virginia, Ph No: 4342976833.

## Abstract

Defects in motile cilia cause a range of disorders, including heterotaxy (HTX), congenital heart disease (CHD), and primary ciliary dyskinesia (PCD). Although these conditions often co-occur, the genetic and mechanistic bases for tissue-specific manifestations remain poorly understood. Here, we identify compound heterozygous variants in DAW1, a dynein arm assembly factor, in a proband with HTX and complex congenital heart disease but no clinical signs of PCD. Whole-genome sequencing revealed a maternally inherited canonical splice-site variant (c.648+1G>A) and a paternally inherited missense variant (c.341G>A; p.Arg114Gln), both classified as variants of uncertain significance under ACMG/AMP guidelines. Using Xenopus tropicalis, we show that Daw1 depletion disrupts left–right patterning, cardiac looping, and mucociliary flow, all of which are rescued by wild-type human DAW1. Functional testing of patient alleles showed notable tissue specificity: p.Arg114Gln fully rescued mucociliary flow but did not restore left–right patterning, while the splice-site variant resulted in a complete loss of function in both contexts. These findings closely match the proband’s clinical phenotype and provide strong functional evidence to support reclassifying c.648+1G>A as pathogenic and p.Arg114Gln as a context-dependent hypomorphic allele. This study establishes functional criteria for interpreting DAW1 variants, shows how developmental context clarifies genotype–phenotype relationships, and highlights how in vivo models can support ACMG reclassification of unresolved HTX-related variants.

## INTRODUCTION

Cilia are highly conserved, microtubule-based projections that extend from the plasma membrane into the extracellular space and are critical in vertebrate development and physiology. They can be broadly divided into immotile (primary) cilia, which function in sensory and signaling processes, and motile cilia, which generate directional fluid flow. During embryogenesis, motile monocilia located at the left-right organizer (LRO), also known as the node in mammals and the gastrocoel roof plate (GRP) in frogs, generate a leftward extracellular flow that is essential for establishing left-right (LR) body asymmetry(Hirokawa *et al*, 2006; Kulkarni & Khokha, 2018; McGrath *et al*, 2003; Rao & Kulkarni, 2021; Schweickert *et al*, 2007). Motile cilia are also present on specialized multiciliated cells (MCCs) that generate unidirectional fluid flow in the respiratory tract, cerebral ventricles, and fallopian tubes in mammals(Rao & Kulkarni, 2021; Spassky & Meunier, 2017; Stannard & O’Callaghan, 2006). Defects in ciliary assembly or motility can lead to motile ciliopathies, including primary ciliary dyskinesia (PCD), heterotaxy (HTX) syndrome, and congenital heart disease (CHD)(Fliegauf *et al*, 2007; Sutherland & Ware, 2009; Wallmeier *et al*, 2020).

PCD is a rare disorder characterized by impaired mucociliary clearance, leading to persistent respiratory complications. PCD’s pulmonary effects are variable but significant(Collins *et al*, 2014; Ferkol, 2025; Hannah *et al*, 2022; Horani *et al*, 2025; Leigh *et al*, 2019; Lie & Ferkol, 2007; Zariwala *et al*, 1993). More than 75% of neonates with PCD experience respiratory distress at birth, necessitating oxygen supplementation for extended periods(De Jesus-Rojas *et al*, 2024; Tilley *et al*, 2015; Zariwala *et al*., 1993). Despite these early signs, PCD diagnoses in neonates are rare(Hosie *et al*, 2014; Zariwala *et al*., 1993). As children grow, they commonly develop symptoms like chronic cough, sputum production, and wheezing, often progressing to obstructive lung disease or bronchiectasis(Hosie *et al*., 2014; Lie & Ferkol, 2007; Zariwala *et al*., 1993). Comorbidities associated with PCD are recurrent otosinopulmonary infections and male infertility (Mirra et al., 2017). About half of PCD patients also exhibit situs inversus (mirror-image reversal of internal organs) without significant physiological effects(Kennedy *et al*, 2007). However, around 12% experience HTX, leading to complex congenital heart disease that can be life-threatening(Kaspy *et al*, 2024; Skeik & Jabr, 2011).

HTX arises from disrupted left-right (LR) body patterning, resulting in discordant organ positioning, frequently accompanied by a wide range of complex CHD phenotypes(Sutherland & Ware, 2009). These include atrioventricular septal defects, atrial isomerism, transposition of the great arteries, double-outlet right ventricle, anomalous pulmonary venous return, single ventricle, and left ventricular outflow tract obstruction(Agarwal *et al*, 2021; Saba *et al*, 2022; Tan *et al*, 2007). These anomalies often require early surgical intervention, yet outcomes remain poor, with high neonatal mortality and lifelong cardiovascular morbidity in survivors. Extra-cardiac manifestations such as splenic abnormalities (asplenia or polysplenia), intestinal malrotation, and pulmonary isomerism may further complicate clinical management(Saba *et al*., 2022). Notably, a substantial portion of patients with HTX also present with respiratory complications resembling PCD(Brueckner, 2007; Harden *et al*, 2014; Kothari, 2014; Nakhleh *et al*, 2012; Swisher *et al*, 2011), indicating a shared genetic basis. Indeed, several genes involved in ciliary function, including dynein heavy and intermediate chains (e.g., DNAH5, DNAH11, DNAI1, DNAI2), structural regulators (e.g., CCDC39, CCDC40), and transcription factors such as FOXJ1, have dual roles in both mucociliary clearance and LR patterning, illustrating how a single variant can manifest as both PCD and HTX(Barber *et al*, 2023; Blanchon *et al*, 2012; Leigh *et al*., 2019; Leslie *et al*, 2022; Raidt *et al*, 2024; Stauber *et al*, 2017; Wallmeier *et al*, 2019; Wilken *et al*, 2024).

Dynein arms are essential for ciliary motility: outer dynein arms (ODAs) generate propulsive force and control beat frequency, while inner dynein arms (IDAs) adjust waveforms and bending(Dai *et al*, 2018; Klena & Pigino, 2022; Walton *et al*, 2023). Proper ODA assembly involves cytoplasmic preassembly and transport into the ciliary axoneme, facilitated by intraflagellar transport (IFT) proteins and specific adaptor molecules(Bearce *et al*, 2022; Klena & Pigino, 2022). Defects in ODA assembly account for the majority of cases in which PCD co-occurs with HTX(Barber *et al*., 2023; Horani *et al*., 2025; Kennedy *et al*., 2007; Raidt *et al*., 2024). Genetic studies in animal models revealed that the protein DAW1 is crucial for the structure and function of motile cilia(Bearce *et al*., 2022; Gao *et al*, 2010; Lesko & Rouhana, 2020; Leslie *et al*., 2022; Solomon *et al*, 2017). In Chlamydomonas, DAW1 (ODA16) interacts with IFT46 to mediate ODA transport, although it remains unclear whether mammalian DAW1 functions similarly remains unclear(Bearce *et al*., 2022; Gao *et al*., 2010). In animal models, the loss of DAW1 disrupts ODA assembly, leading to laterality defects(Gao *et al*., 2010). In humans, predicted pathogenic DAW1 variants have been associated with HTX, CHD, and chronic respiratory symptoms(Leslie *et al*., 2022).

Here, we report on a proband with HTX and complex CHD with no evidence of PCD, who carries compound heterozygous DAW1 variants: a paternally inherited missense mutation (c.341G>A; p.Arg114Gln) and a maternally inherited splice-site mutation (c.648+1G>A). Both variants are classified as variants of uncertain significance (VUS) and are rare in population databases such as gnomAD. To investigate their pathogenicity, we used *Xenopus tropicalis* as a model system to examine their effects on L-R patterning, cardiac looping, and mucociliary flow. This study provides experimental evidence that patient-derived DAW1 mutations disrupt laterality development and highlight tissue-specific requirements of DAW1 in human disease.

## RESULTS

### Clinical presentation of the patient

The proband is a male infant born at 37 + 6 weeks of gestation to a mother with type 2 diabetes, treated with insulin during pregnancy. The pregnancy was notable for prenatal concerns of congenital heart disease, and the infant was transferred to the NICU after delivery. Echocardiography demonstrated a double-outlet right ventricle with D-malposed great arteries, a large subpulmonic VSD with inlet extension, a posterior muscular VSD, moderate PDA, and a small secundum ASD/PFO with combined left-to-right shunting. RV systolic function was qualitatively normal, and LV size and function were preserved. Cardiac MRI confirmed these findings and additionally demonstrated a thin membranous structure attached to the interventricular septum and directed toward the tricuspid valve. The infant underwent balloon atrioseptostomy, followed by pulmonary artery band placement. Head ultrasound revealed a left grade 1 germinal matrix hemorrhage without ventriculomegaly, and the abdominal ultrasound was normal. On physical examination, the patient was nondysmorphic with no extracardiac malformations.

Family history was negative for consanguinity. Given the proband’s presentation with complex congenital heart disease and family history, whole genome sequencing (WGS) with parental samples was pursued through GeneDx after informed consent.

### Whole Genome Sequence analysis and findings

Analysis of variants identified from whole-genome sequencing (WGS) data and prioritized using Genomiser(Smedley *et al*, 2016) identified DAW1 as the top candidate gene, with a phenotype similarity score of 0.683, driven by concordance between the proband’s clinical features and *Primary ciliary dyskinesia 52*. Two rare compound heterozygous variants in DAW1 were detected:

DAW1 (ENST00000309931.3): c.648+1G>A, p.? (rs927376980). This splice donor variant has not been previously reported in ClinVar and was classified as a variant of uncertain significance (VUS) by both Genomiser and Exomiser according to ACMG/AMP guidelines(Richards *et al*, 2015) (Exomiser ACMG: *UNCERTAIN_SIGNIFICANCE* [PM2_Supporting, PP4]). The variant is extremely rare in population databases, with a maximum allele frequency of 1.33 × 10⁻⁵ observed in individuals of African/African American ancestry. LOFTEE(Karczewski *et al*, 2020) predicts this variant to be a high-confidence loss-of-function allele. Splice prediction analyses indicate that the most likely consequence is exon 7 skipping, resulting in an in-frame deletion (p.Val181_Arg216del). A less likely alternative outcome is the activation of a cryptic splice donor site approximately 459 bp downstream, potentially leading to premature truncation.

DAW1 (ENST00000309931.3): c.341G>A, p.(Arg114Gln) (rs759511456). This missense variant has not been reported in ClinVar and was also classified as a VUS by Genomiser and Exomiser (Exomiser ACMG: *UNCERTAIN_SIGNIFICANCE* [PP4]). The variant is rare in population databases, with a maximum allele frequency of 5.13 × 10⁻⁴ observed in East Asian ancestry. The substitution has a Phred-scaled CADD(Rentzsch *et al*, 2019) score of 28, placing it among the top 0.16% of predicted deleterious variants in the human genome.

Both variants segregated in the proband in a compound heterozygous configuration, consistent with autosomal recessive inheritance, and were therefore prioritized for downstream structural modeling and functional interpretation. Additional candidate genes with non-zero phenotype similarity scores were identified but were considered less likely contributors to the phenotype, as these genes were predicted to act in an autosomal-dominant manner, and neither parent exhibited overlapping clinical features. The secondary candidates included RYR1 (phenotype score 0.522; similarity to King–Denborough syndrome), KAT8 (phenotype score 0.522; similarity to Li--Ghorgani--Weisz--Hubshman syndrome), and CCDC22 (phenotype score 0.538; similarity to Ritscher--Schinzel syndrome type 2).

### In silico predictions and AlphaFold modeling

We assessed the predicted pathogenicity of the compound heterozygous DAW1 variants using multiple *in silico* algorithms and AlphaFold modeling. For the missense variant c.341G>A (p.Arg114Gln; R114Q) inherited from the father, Arginine is highly conserved across taxa (Fig. 1). Computational predictors yielded inconsistent results, producing an overall classification of uncertain significance (Table 1). Revel (0.39), SIFT (0.004), FATHMM (0.1), MetaLR (0.27), and PrimateAI (0.54) indicated an uncertain or benign effect (Table 1). The MetaLR logistic regression–based ensemble score, which integrates ten independent predictors (SIFT, PolyPhen-2 HDIV, PolyPhen-2 HVAR, GERP++, MutationTaster, MutationAssessor, FATHMM, LRT, SiPhy, PhyloP) with population allele frequencies, also supported a benign effect (0.27). In contrast, AlphaMissense (0.78), MutationAssessor (2.75), MutationTaster (1), and DANN (1) predicted a deleterious impact. AlphaFold structural modeling showed no discernible difference relative to the wild-type protein (Fig. 1). Overall, although some algorithms showed partial support for a deleterious effect, these findings suggest that R114Q remains of uncertain pathogenicity.

**Figure 1:**
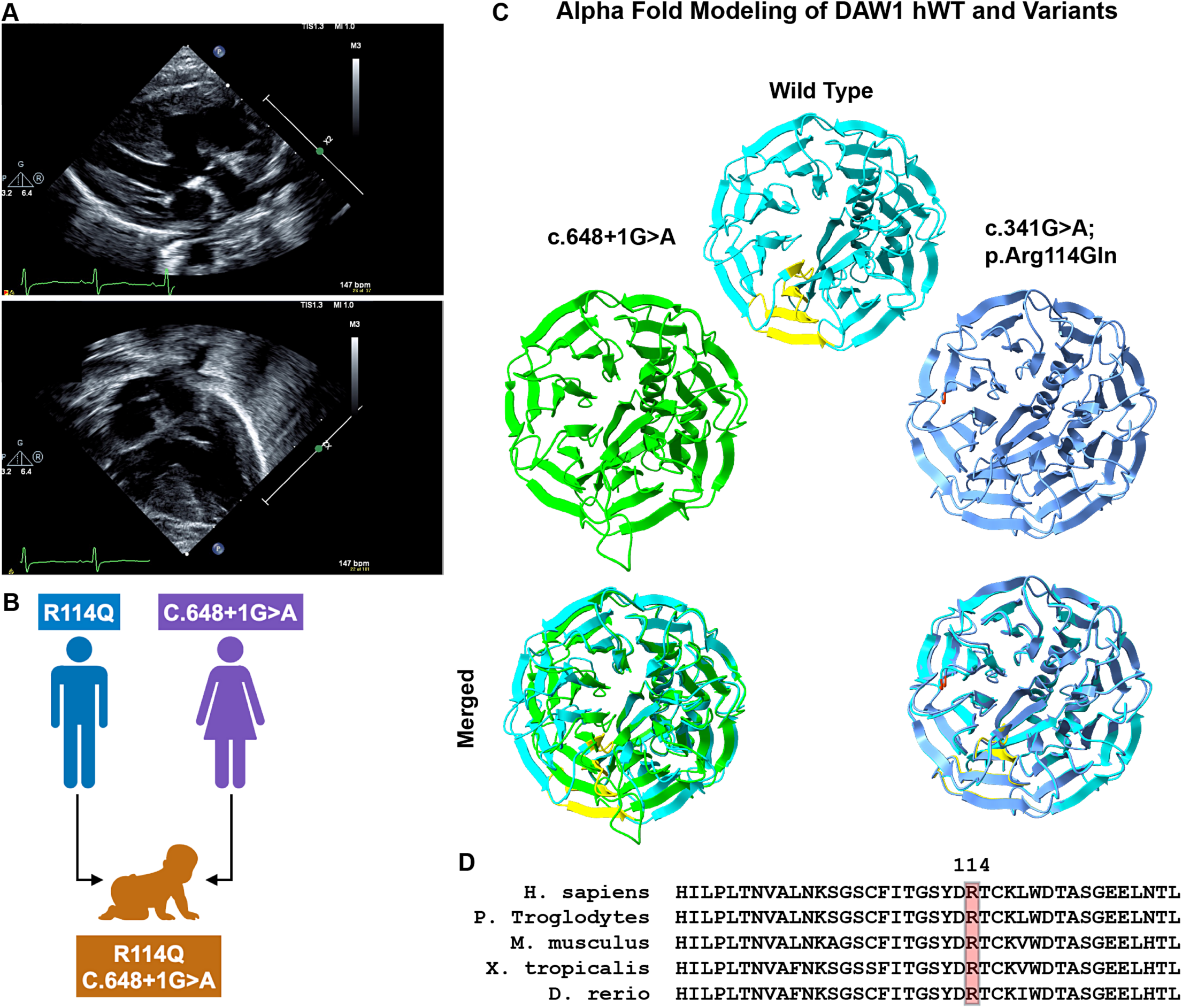
Clinical presentations of the patient and *in silico* predictions. A. Top image: Long-axis view demonstrates DORV with both great arteries arising from the RV and a mitral/aortic discontinuity. What appeared to be a subpulmonic VSD did not have flow across the VSD by echo, and this was confirmed angiographically. Bottom image: Apical four-chamber view demonstrates a posterior, high-muscular VSD that was the outlet of flow from the LV, and at the second surgery, was successfully baffled to the neo-aortic valve. This, in combination with the arterial switch, resulted in a biventricular circulation. B. Graphical representation of compound heterozygous inheritance. The patient has compound heterozygous variants in *DAW1*, including a paternally inherited c.341G>A (p.R114Q) variant and a maternally inherited c.648+1G>A (p.?) variant. Consistent with autosomal recessive inheritance, both parents are unaffected carriers, with a 25% recurrence risk in each subsequent pregnancy. C. AlphaFold-predicted structure of wild-type DAW1 (cyan) compared with the splice-site (green) and R114Q (blue) mutant proteins. Residues 181–216, which are absent in the splice-site mutant, are highlighted in yellow. The R114Q substitution is indicated in orange. D. Multiple sequence alignment of DAW1 across representative vertebrate species highlighting conservation surrounding the R114 residue.

**Table 1:**
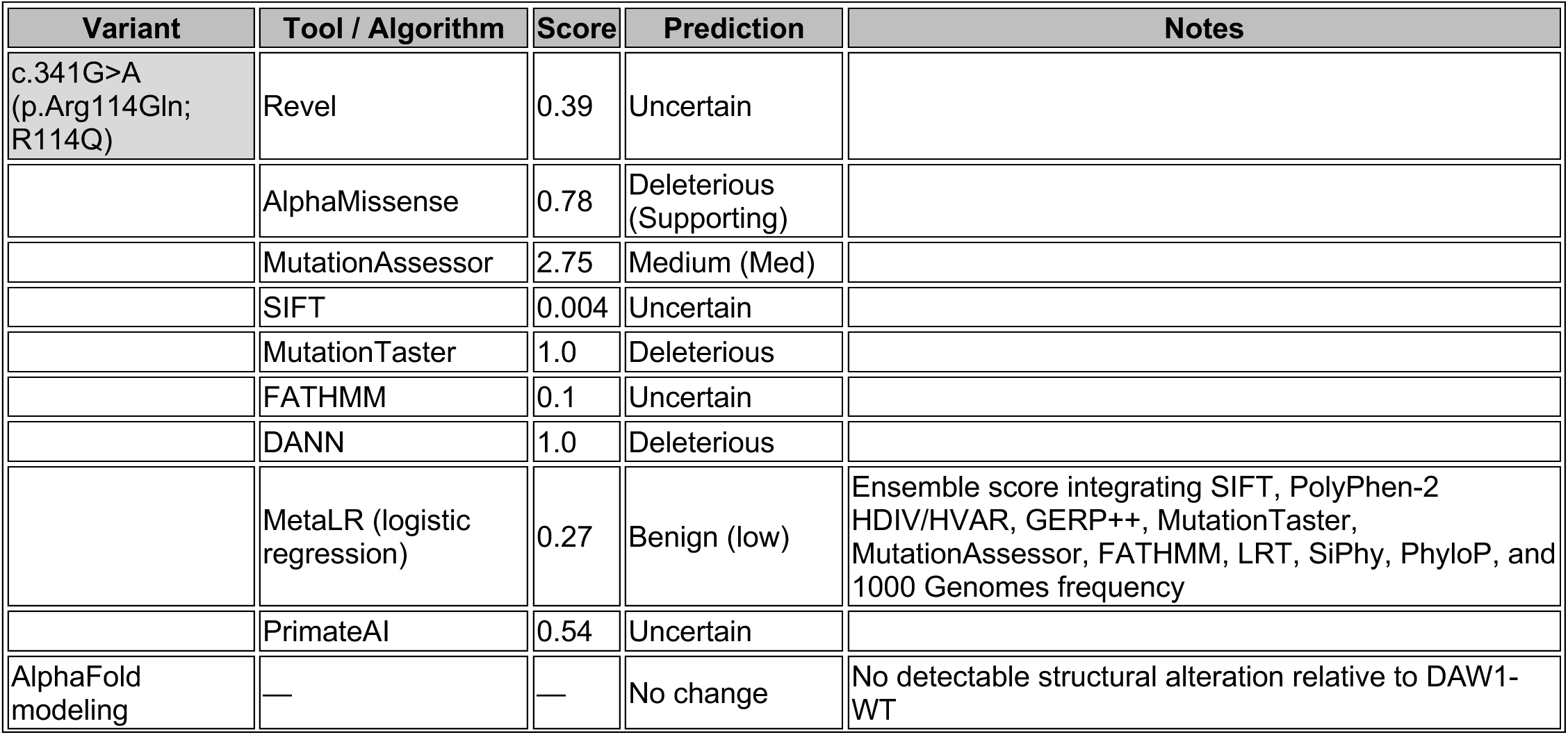
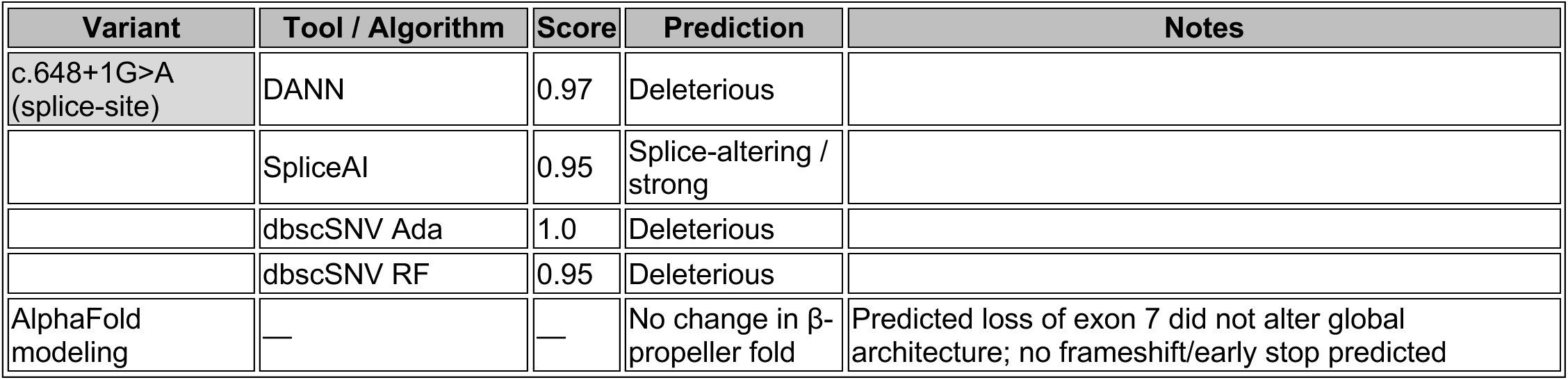
Scores are reported as raw outputs from each tool, with corresponding categorical interpretations (benign/uncertain/deleterious) provided according to the authors’ guidelines. AlphaFold structural predictions were compared with those of the wild-type DAW1 model.

In contrast, the canonical splice-site variant c.648+1G>A, inherited from the mother, was consistently predicted to be deleterious. DANN (0.97), SpliceAI (0.95), dbscSNV Ada (1), and dbscSNV RF (0.95) all strongly suggest splice disruption (Table 1). Exon 7 skipping was predicted, although AlphaFold modeling of the resulting transcript did not show major perturbation of the β-propeller structure of DAW1 (Fig. 1). This may be because the splicing of exon 7 does not change the reading frame or introduce a premature stop codon, despite being expected to impair splicing.

Together, these findings indicate that the R114Q missense variant remains a variant of uncertain significance, while the splice-site variant c.648+1G>A is strongly predicted to disrupt normal DAW1 splicing. Detailed scores and categorical outputs from all in silico prediction tools are provided in Table 1.

### *Daw1* knockdown *in vivo* affects left-right patterning and cilia motility in *X. tropicalis* embryos

HTX was the predominant phenotype in the patient. Therefore, we investigated the effects of a *daw1* knockdown on LR patterning in *X. tropicalis* embryos using a morpholino oligonucleotide (MO). We performed the whole-mount *in situ* hybridization for *pitx2*, a marker of LR asymmetry at embryonic Stage 28(Kulkarni *et al*, 2024). *pitx2* is normally expressed on the left side of the lateral plate mesoderm downstream of cilia-mediated leftward flow in the LRO(Ryan *et al*, 1998). While control embryos showed normal left-sided expression, Daw1 morphants showed significantly more abnormal *pitx2* expression (right-sided, bilateral, or absent) (Fig. 2A, B). To further evaluate LR patterning and cardiac development, we assessed cardiac looping at embryonic stage 48 (72-96 hours post fertilization, hpf)(Arrigo *et al*, 2025; Blum *et al*, 2009). A D-loop of the outflow tract (left to right, *situs solitus*) was classified as normal, whereas an L-loop (right to left, *situs inversus*) and an A-configuration (straight back, HTX) were classified as abnormal. Daw1 depletion resulted in a significant increase in heart-looping defects compared with controls (Fig 2C, D). Together, these findings suggest that the absence of Daw1 disrupts LRO flow, as indicated by abnormal *pitx2* expression, and leads to defects in laterality and organ morphogenesis, as demonstrated by aberrant heart looping.

**Figure 2:**
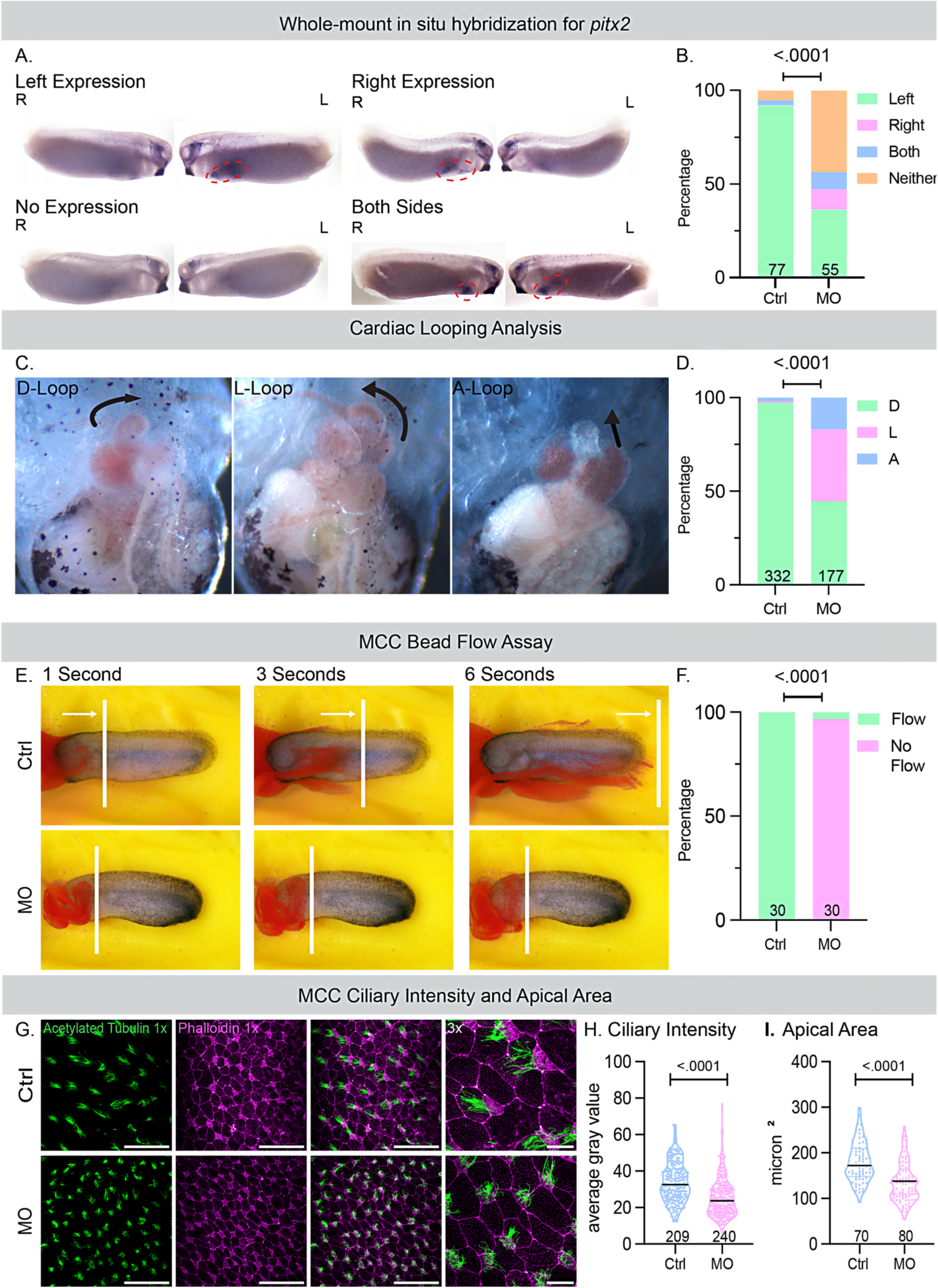
DAW1 is essential for LR patterning and mucociliary flow in the *Xenopus* embryos. A. Whole-mount *in situ* hybridization for *pitx2*, where normal expression appears only on the left side of the embryo, can be compared to abnormal expression, which may be present only on the right side, bilaterally, or absent altogether. The embryos shown were at Stage 28. B. Quantification of expression in wild type (Ctrl) and 20ng *daw1* MO-injected embryos at Stage 28. The Ctrl embryos (n=77, N=3) showed left expression (green) of *pitx2* in 71 embryos, which was considered normal. Abnormal expression included 2 embryos with bilateral staining (blue), 4 with no staining (orange), and no embryos had right-sided expression (pink). The MO embryos (n=55, N=3) had 20 with normal expression and 35 with abnormal expression, including 6 with right-sided expression, 24 with no expression, and 5 with bilateral expression. A Chi-Square test comparing Ctrl and MO embryos revealed a significant difference in *pitx2* expression (pf < .0001). C. Cardiac looping categories, where a D-loop is considered normal, and the A-loop and L-loop formations are considered abnormal. D. Quantification of the three loop types between Ctrl embryos and MO embryos. The Ctrl embryos (n=332, N=3) had 323 hearts with normal cardiac D looping (shown in green) and 9 with abnormal looping (3 in the L configuration, shown in pink, and 6 in the A configuration, shown in blue), while the MO embryos (n=177, N=3) had 79 hearts with normal cardiac looping and 98 with abnormal looping (68 in the L configuration and 30 in the A configuration). A Chi-Square test comparing Ctrl and MO embryos showed a significant difference in cardiac looping (p < .0001). E. Still frames from videos at 1 second, 3 seconds, and 6 seconds show the movement of latex microspheres (beads) over the epidermis of Ctrl embryos and MO embryos. The position of the furthest bead throughout the videos is marked by a white vertical line. F. The number of Ctrl and MO injected embryos showing the movement of beads as “Flow” and no movement as “No Flow” is quantified. The Ctrl embryos (n=30, N=3) all exhibited flow, while the MO embryos (n=30, N=3), 29 showed no flow and 1 showed flow. A Chi-Square analysis revealed a significant difference in the presence of flow between Ctrl and MO embryos (p < .0001). G. Confocal imaging of the epidermis stained for acetylated tubulin (green) to visualize cilia and phalloidin (magenta) to visualize filamentous actin for cell boundaries and MCCs. The first three images are 1x images, with a scale bar of 100 μm, showing, respectively, only acetylated tubulin staining, only phalloidin staining, and a composite of the two. The fourth image from the left is a 3x composite image with a scale bar of 20 μm. H. Quantification of ciliary intensity in Ctrl and MO embryos. For quantification, approximately 15 cells were analyzed, with 14 control embryos and 16 MO embryos across three trials. A Welch’s t-tes test revealed a significant difference in ciliary intensity between the control embryos (n=209, N=3) and MO embryos (n=240, N=3) (p <.0001). I. For quantifying the apical area in Ctrl and MO embryos, five cells were measured per embryo, with 14 control embryos and 16 MO embryos across three trials. A Welch’s t-test showed a significant difference in apical area between the control group (n=70, N=3) and MO embryos (n=80, N=3) (p < .0001).

Beyond its role in LR development, motile cilia are also vital for airway mucociliary clearance. Indeed, patients with CHD, and especially HTX, often suffer from chronic respiratory issues due to ciliary dysfunction(Harden *et al*., 2014; Nakhleh *et al*., 2012; Swisher *et al*., 2011). A previous study with patients with DAW1 variants described chronic respiratory dysfunction, suggesting that DAW1 may play a significant role in mucociliary clearance(Leslie *et al*., 2022). We therefore examined the function of Daw1 in the multiciliated cells (MCCs) of the *X. tropicalis* embryonic epidermis, a well-established *in vivo* system for analyzing mucociliary flow.

We visualized mucociliary flow by adding fluorescent latex microspheres (beads) to the culture medium for time-lapse imaging of bead movemen(Kulkarni *et al*, 2018). In control embryos, fluorescent beads placed on the anterior of the embryo were rapidly transported toward the posterior, reflecting coordinated ciliary beating. Daw1 depletion with MO led to a significant loss of cilia-generated fluid flow, indicating either a significant loss of cilia motility or assembly (Fig. 2E). To test these possibilities, we performed immunofluorescence using acetylated tubulin to label the ciliary axoneme and phalloidin to label F-actin and the apical size of the cells. Compared with controls, Daw1-depleted embryos exhibited a small but significant reduction in the apical surface area of MCCs and lower normalized ciliary fluorescence intensity, consistent with impaired ciliogenesis (Fig. 2G-I). These results demonstrate that Daw1 is essential for both L-R patterning and mucociliary clearance function *in vivo*.

### Wild-type human DAW1 rescue of left-right patterning and mucociliary flow

To test the specificity of the MO and establish the function of wild-type (WT) - human DAW1 in *Xenopus*, we performed a rescue of the LR patterning and loss of mucociliary flow phenotypes. First, we co-injected an RNA construct encoding WT-hDAW1 fused to an MStayGold (msg) fluorescent tag at the C-terminus with Membrane RFP RNA to label the ciliary axonemes of MCCs in control embryos and assessed its localization. WT-hDAW1-msg localized to the basal bodies and ciliary axonemes of MCCs (Fig. 3A). We also expressed the DNA of WT-hDAW1-msg and observed the same localization as DNA (Fig. 3B). Next, we co-injected the WT-hDAW1-msg with *daw1* MO to assess the rescue. WT-hDAW1-msg significantly rescued both mucociliary flow and heart-looping (LR patterning) phenotypes relative to MO embryos lacking WT-hDAW1-msg (Fig. 3C, D). These results laid the foundation for testing the function of *DAW1* variants in LR patterning and mucociliary flow.

**Figure 3:**
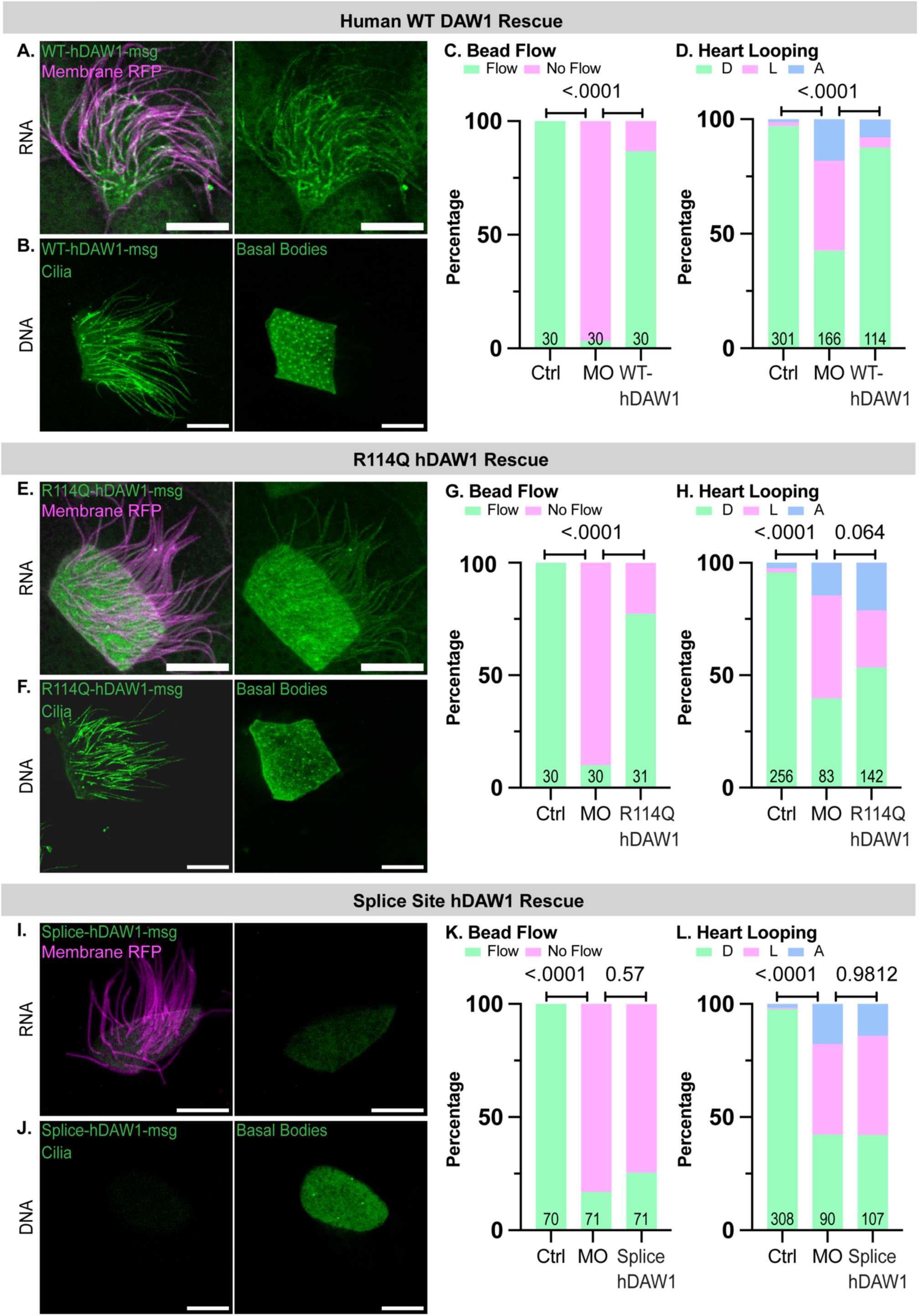
DAW1 variants show tissue-specific pathogenic effects. All Scale Bars are 10uM long A. The left panel shows a composite image of both Membrane RFP (ciliary marker) and WT-hDAW1-msg constructs in an MCC, while the right panel shows only the WT-hDAW1-msg expression. B. Beneath the RNA expression, WT-hDAW1-msg DNA plasmid localization is shown at the ciliary level in the left panel and the basal level in the right panel. C. Quantification of bead flow in uninjected control embryos, embryos injected with *daw1* MO, and embryos injected with both MO and WT-hDAW1-msg (referred to as WT-hDAW1 in the chart). All control embryos showed flow (displayed in green) (n=30, N=3). MO embryos had 29 with no flow (pink) and 1 with flow (n=30, N=3). WT-hDAW1 coinjected embryos had 26 with flow and 4 without flow (n=30, N=3). The Chi-Square Test revealed significant differences between control and MO embryos (p < .0001) and between MO and WT-hDAW1 embryos (p < .0001). D. Quantification of heart looping in uninjected control embryos, embryos injected with MO, and embryos injected with both MO and WT-hDAW1-msg. The control embryos (n=301, N=3) had 292 D loops, with normal hearts (shown in green), and 9 embryos showed abnormal configurations: 5 with L loops (shown in pink) and 4 with A loops (shown in blue). The MO embryos (n=166, N=3) included 71 with normal D loops and 95 with abnormal configurations: 65 with L loops and 30 with A loops. The WT-hDAW1-injected embryos (n=114, N=3) had 100 with normal D loops and 14 with abnormal configurations: 5 with L loops and 9 with A loops. A Chi-Square test showed significant difference in heart looping between control and MO embryos (p = <.0001), and also between MO and WT-hDAW1-injected embryos (p <.0001). E. The left panel shows a composite image of the expression of both R114Q-hDAW1-msg RNA and Membrane RFP RNA constructs in an MCC, and the right panel shows only the R114Q-hDAW1-msg expression. F. The ciliary localization of R114Q-hDAW1-msg DNA plasmid is shown in the left panel, and the basal localization is shown in the right panel. G. Quantification of bead flow in uninjected control embryos, embryos injected with *daw1* MO, and embryos injected with both MO and R114Q-hDAW1-msg (referred to as R114Q-hDAW1 in the figure). All control embryos showed flow (depicted in green) (n=30, N=3), while MO embryos (n=30, N=3) had 27 without flow (shown in pink) and 3 with flow. The R114Q-hDAW1-msg embryos (n=31, N=3) had 24 with flow and 7 without flow. Yates’ Chi-Square test revealed a significant difference between the control and MO embryos (p <.0001). A Chi-Square test also showed a significant difference between the MO and R114Q-hDAW1-msg embryos (p < .0001), indicating successful flow rescue with the R114Q-mutated DAW1 protein. H. Quantification of heart looping in uninjected control embryos, embryos injected with MO, and embryos injected with both MO and R114Q-hDAW1-msg. The control embryos (n= 256, N=3) had 245 D loops, which are normal hearts (shown in green), and a total of 11 embryos exhibited abnormal configurations, with 5 having L loops (shown in pink) and 6 having A loops (shown in blue). The MO embryos (n=83, N=3) included 33 with normal D loops, and a total of 50 with abnormal configurations, with 38 being L loops and 12 A loops. The R114Q-hDAW1-msg embryos (n=142, N=3) showed 76 with normal D loops and 66 with abnormal configurations, including 36 with L loops and 30 with A loops. A Chi-Square test revealed a significant difference in the number of normal versus abnormal embryos between the control and the MO groups (p < .0001). No significant difference was found between the MO and R114Q-hDAW1-msg groups (p = 0.064). I. The left panel displays a composite image of the expression of both RNA constructs in an MCC, while the right panel shows only the Splice-hDAW1-msg expression. J. Splice-hDAW1-msg DNA plasmid localizes to ciliary axonemes and basal bodies in MCCs. The ciliary level is shown in the left panel, and the basal level is shown in the right panel. K. Quantification of bead flow in uninjected control embryos, embryos injected with *daw1* MO, and embryos injected with both MO and Splice-hDAW1-msg (referred to as Splice-hDAW1 in the figure). All control embryos showed flow (indicated in green) (n=70, N=7), while MO embryos (n=71, N=7) had 59 with no flow (shown in pink) and 12 with flow. The Splice-hDAW1-msg injected embryos (n=71, N=7) had 18 with flow and 53 with no flow. Yates’ Chi-Square test revealed a significant difference between control and MO embryos (p <.0001). However, Chi-Square did not indicate a significant difference between MO and Splice-hDAW1-msg embryos (p = 0.57). L. Quantification of heart looping in uninjected control embryos, embryos injected with MO, and embryos injected with both MO and Splice-hDAW1-msg. The control embryos (n=308, N=5) mostly had 301 D loops, which are normal hearts (shown in green), while a total of 7 embryos had abnormal configurations: 2 with L loops (shown in pink) and 5 with A loops (shown in blue). The MO embryos (n=90, N=5) included 38 with normal D loops, and 52 with abnormal configurations: 36 with L loops and 16 with A loops. The Splice-hDAW1-msg embryos (n=107, N=5) had 45 with normal D loops and 62 with abnormal configurations: 47 with L loops and 15 with A loops. A Chi-Square test showed a significant difference between the control and the MO embryos (p <.0001). The Chi-square test did not show a significant difference between the MO embryos and the Splice-hDAW1-msg (p = 0.9812).

### Context-specific rescue with the missense mutation

Individual mutant constructs tagged with the C-terminus-msg were then generated for each of the patient’s *DAW1* mutations. The paternally inherited missense variant c.341 G>A, p.(Arg114Gln) (R114Q) was first examined. Injections of both RNA and DNA showed that localization of the R114Q protein at the basal bodies and ciliary axoneme of MCCs was similar to WT-hDAW1 (Fig. 3E, F). Given that localization was unaffected, we performed functional assays to assess pathogenicity. At the developmental Stage 28 (24 hpf), R114Q-hDAW1-msg-injected embryos showed improved mucociliary flow relative to MO embryos lacking R114Q-hDAW1-msg (Fig. 3G). To assess LR patterning, we raised the same embryos to developmental stage 48 (72 hpf) to analyze heart looping. Interestingly, LR patterning was not rescued relative to MO embryos lacking WT-hDAW1 (Fig. 3H), suggesting that the R114Q mutation affects cilia function in a tissue-specific context.

### Loss-of-function splice-site variant

A second variant, the maternally inherited splice-site mutation c.648+1 G>A, was predicted to be more disruptive than R114Q due to skipping of entire exon 7 (E7) as a most likely output (Fig. 1). Injection of both RNA and DNA of Splice-hDAW1-msg showed a consistent loss of localization at the basal bodies and ciliary axonemes of MCCs suggesting complete loss of function (LOF) (Fig. 3I, J). To functionally confirm our localization results, we examined both LR patterning and mucociliary flow using the rescue experiments described above for the missense mutation. As expected, the Splice-hDAW1-msg did not rescue either phenotype, confirming the complete LOF in both LR patterning and mucociliary clearance contexts.

## DISCUSSION

Cilia are essential organelles that regulate fluid movement during development and homeostasis. Motile cilia in the LRO generate leftward flow needed for proper LR patterning during embryogenesis, while motile cilia of MCCs mediate mucociliary clearance in the respiratory tract and fluid circulation in other organ systems. Defects in these processes can cause a wide range of motile ciliopathies, including PCD, HTX, and CHD(Wallmeier *et al*., 2020). Here, we identify compound heterozygous DAW1 variants in a patient presenting with HTX and complex CHD, but notably did not exhibit features of PCD. This phenotype, CHD/HTX in the absence of respiratory disease, was precisely recapitulated in *Xenopus tropicalis*, providing strong experimental validation of the genetic findings.

*In silico* predictors failed to reach consensus on the R114Q missense variant, with results ranging from benign to deleterious. AlphaFold modeling similarly showed no structural disruption, highlighting the limitations of current computational tools in determining the pathogenicity of subtle variants. Therefore, functional assays were crucial. Notably, R114Q displayed a context-specific effect: it completely restored mucociliary flow in MCCs but did not rescue LR patterning (heart looping) defects, closely reflecting the proband’s phenotype of HTX without chronic respiratory disease. These findings illustrate that DAW1 function can diverge across ciliary subtypes and that single amino acid substitutions can selectively affect LRO cilia without impairing MCC function. Such specificity cannot currently be captured by in silico prediction models, emphasizing the importance of functional validation in relevant developmental contexts.

In contrast, the splice-site variant c.648+1G>A functions as a complete loss-of-function allele. Both in silico splicing tools and functional assays indicated exon 7 skipping, loss of proper localization, and failure to rescue either L–R patterning or MCC flow. These findings confirm c.648+1G>A as a deleterious variant and demonstrate that compound heterozygosity for R114Q and c.648+1G>A can fully explain the proband’s phenotype. Notably, the *Xenopus* model mirrored the clinical presentation, underscoring its utility as a rapid and reliable model for analyzing genotype–phenotype relationships in ciliopathies. Together, these data support reclassification of c.648+1G>A as pathogenic and identify p.Arg114Gln as a context-dependent hypomorphic allele whose effects are not detected by current in silico or ACMG/AMP guidelines (Table 2).

**Table 2.** ACMG/AMP evidence supporting interpretation of DAW1 variants.

| Variant | Evidence type | ACMG code | Strength | Evidence summary |
| --- | --- | --- | --- | --- |
| c.648+1G>A | Functional assay | PS3 | Strong | Complete loss of localization and failure to rescue LR patterning and MCC flow in *Xenopus* |
| c.648+1G>A | In trans with hypomorphic allele | PM3 | Moderate | Compound heterozygous in affected proband |
| c.648+1G>A | Phenotype match | PP4 | Supporting | HTX and CHD without PCD consistent with DAW1 spectrum |
| c.648+1G>A | Allele frequency | PM2 | Supporting | Maximum of $1.33 \times 10^{-5}$ observed in individuals of African/African American ancestry |
| p.Arg114Gln | Functional assay | PS3 | Supporting | Selective rescue of MCC flow but not LR patterning |
| p.Arg114Gln | Phenotype match | PP4 | Supporting | Explains laterality defect without respiratory disease |
| p.Arg114Gln | Allele frequency | PM2 | Supporting | Maximum of $5.13 \times 10^{-4}$ observed in East Asian ancestry |

Our results also build upon and expand previous reports of DAW1-related disease. In the largest published series, Leslie et al. described several families with different DAW1 genotypes and varying effects on laterality and respiratory features(Leslie *et al*., 2022). For example, homozygous p.(Asn143Asp) variants were associated with *situs inversus* without respiratory symptoms in two patients, whereas another patient exhibited respiratory symptoms without laterality defects. A homozygous p.(Trp119∗) nonsense variant resulted in complex CHD with situs ambiguous, a transverse liver, and a right-sided spleen, but no respiratory symptoms. Compound heterozygous variants p.(Leu66∗)/p.(Trp372Cys) and homozygous p.(Ser364Thr) were both reported in patients with complex CHD resembling HTX, although respiratory involvement was not specified. Functional studies in zebrafish further supported allele-specific effects, with p.(Asn143Asp) and p.(Ser364Thr) showing complete loss of function, while p.(Trp372Cys) was a hypomorph based on the rescue of cardiac looping and cilia motility in Kupffer’s vesicle(Leslie *et al*., 2022).

In this context, our study offers the first direct evidence that patient-derived DAW1 variants can differentially affect LRO and multiciliated cilia in *X. tropicalis*, explaining why the proband exhibited HTX and CHD without respiratory disease. The R114Q allele acted as a context-specific hypomorph (affecting LR patterning but not mucociliary flow), whereas the splice-site variant resulted in complete loss of function, together creating the compound heterozygous state observed clinically. This accurate replication of the human phenotype highlights both the tissue-specific functions of DAW1 and the value of *Xenopus* as a translational model.

While our findings demonstrate functional effects for DAW1 variants, several aspects warrant further exploration. The *Xenopus tropicalis* model provides a rapid and robust system for studying DAW1 function *in vivo*; however, additional validation using human respiratory or cardiac cell models would enhance its translational significance. Similarly, direct RNA analysis of patient-derived tissue is necessary to confirm the predicted exon 7 skipping due to the splice-site variant. Lastly, given the family history of respiratory and auditory features, the influence of genetic background and modifier alleles warrants further investigation. Future research involving patient iPSCs, human airway cultures, and larger clinical cohorts will help clarify the phenotypic spectrum and genotype–phenotype correlations of DAW1 variants.

In summary, this study provides functional validation for compound heterozygous DAW1 variants in a patient with HTX and complex CHD. The ability of *Xenopus tropicalis* to reproduce this precise phenotype underscores its unique capacity to link patient genotypes to mechanistic outcomes. We demonstrate that relying solely on *in silico* tools is insufficient to predict the pathogenicity of DAW1 variants, and that developmental models are crucial for revealing context-specific requirements of ciliary assembly factors.

## METHODS

### IRB protocol

The family was recruited under the IRB protocol HSR210285.

### Sequence analysis

Sequence reads were processed using a Nextflow workflow (https://github.com/aakrosh/PedigreeVarFlow). As part of the workflow, genome reads were aligned to the GRCh38 reference genome with BWA-MEM(Li, 2013) (v. 0.7.19-r1273), and SAMBLASTER (v. 0.1.26)(Faust & Hall, 2014) was used to flag putative PCR duplicates and add MC/MQ tags to paired-end alignments. Resulting SAM files were converted to BAM format and coordinate-sorted using samtools (v. 1.21.42)(Danecek *et al*, 2021). Alignment statistics and quality metrics were generated with alignstats (v. 0.11, https://github.com/jfarek/alignstats).

Variants were called using FreeBayes(G, 2012) (v. 1.3.9 with default parameters) and filtered with bcftools (v. 1.21) to remove low-confidence calls. We applied the following filters to retain variants with strong read support: QUAL>1 && QUAL/INFO/AO>10 && INFO/SAF>0 && INFO/SAR>0 && INFO/RPR>1 && INFO/RPL>1. Variants overlapping known problematic genomic regions were flagged during annotation. Variant-level quality metrics were summarized using Variant QC(Yan *et al*, 2019).

Variants were left-aligned and normalized using bcftools prior to downstream analysis. Candidate variants were prioritized with Genomiser using recommended best practices (REVEL, MVP, AlphaMissense, and SpliceAI variant pathogenicity prediction sources and human-only hiPHIVE gene:phenotype associations, ClinVar whitelist, inheritance filters) and the following human phenotype ontology terms: *Double outlet right ventricle, Dextrotransposition of the great arteries,* and *Subarterial ventricular septal defect(Cooperstein et al, 2025)*. In parallel, variants were annotated using AutoGVP(Kim *et al*, 2024), which integrates germline pathogenicity data from ClinVar and applies ACMG guideline-based classifications using a modified version of InterVar.

### Sequence Alignment and Alphafold modelling

Multiple sequence alignment was performed using the MultAlin web server. Sequences corresponding to residues 91–130 were obtained from human (UniProt: Q8N136-1), *Xenopus tropicalis* (UniProt: Q6P2Y2), *Pan troglodytes* (UniProt: H2QJJ9), mouse (UniProt: D3Z7A5), and Zebrafish (*Danio rerio*, UniProt: Q1LV15). Predicted structural models of human DAW1-WT (UniProt: Q8N136-1), R114Q, and the splice-site mutant were generated using AlphaFold3. Structural visualization and annotation were performed in UCSF ChimeraX.

### Animal Husbandry and microinjections

*Xenopus tropicalis* were bred, housed, and cared for in our aquatics facility according to established protocols (ACUC# 4295) that were approved by the University of Virginia Institutional Animal Care and Use Committee (IACUC). Embryos needed for experiments were produced by *in vitro* fertilization according to previously established protocols(Kulkarni *et al*, 2021; Kulkarni *et al*., 2024). Briefly, testes are removed from the male and crushed in 1xMBS + 0.2%BSA and added to eggs obtained from hCG-injected female frogs. The eggs and sperm are incubated for 3 minutes before being flooded with 0.1x MBS (pH 7.8–8) for 10 minutes. Fertilized eggs were then dejellied using 3% Cysteine in 1/9MR (pH 7.8–8) for 6 minutes. Embryos were then washed using 0.1xMBS and used for microinjections in 1/9MR+Gentamicin. Staging of Xenopus tadpoles was as previously described(Nieuwkoop, 1994).

### Cloning and mRNA synthesis

The full-length human DAW1 (NM 178821.3) and mStayGold was subcloned into the pCS2+ vector using PCR amplification using Gibson assembly to generate DAW1-mStayGold. The primers used for PCR are provided in the resource table. The missense and splice-site variants were generated by site-directed mutagenesis using the WT plasmid as a template. For mRNA synthesis, the plasmids were linearized with NotI and used as templates. Capped mRNAs were synthesized *in vitro* using the mMessage and mMachine SP6 transcription kit following the manufacturer’s instructions.

### Morpholino and mRNA microinjections

Morpholino oligonucleotides (MO) or mRNA were injected into one-cell or four-cell embryos as described previously(Narayanan *et al*, 2025). For most experiments, the translation-blocking MO for *Daw1* (AAGGAATCGCTTTAGCCGCATCGTG) was injected at 20 ng at the one-cell stage, along with Oregon green 488-labelled Dextran (10 kDa, non-fixable), a tracer for all flow and heart looping trials (described below). The DAW1 mRNAs were injected at 200pg in the one-cell stage. For some experiments, the plasmid DNA (WT-hDAW1-msg and the variants) was injected at a 100 pg concentration with the membrane RFP mRNA (100 pg) in one of the 4 blastomere. Post-injection, the embryos were allowed to develop to the appropriate stage for further experiments. For rescue experiments, the embryos were injected with 20ng of DAW1 MOmixed with 200pg of either WT, or variant mRNA.

### Immunofluorescence, image analysis and statistics

Confocal imaging was done on embryos once they reached stage 28 either live or fixed.

For fixation, 4% paraformaldehyde (PFA) was used then the embryos were washed three times with PBST (1× PBS with 0.2% Triton X-100) for 10 min each and then incubated in a blocking solution (3% BSA in PBST) for 1 hour. The primary antibody Mouse Monoclonal Anti-Acetylated α-tubulin was added to the embryos, incubated for 1 hour at room temperature, and washed three times for 10 min each with PBST. Dilutions of the secondary antibody Chicken anti-mouse conjugated to Alexa fluor 488 and the Actin stain Phalloidin in PBST were used to stain embryos for 1 hour. All live imaging was done with Stage 28 embryos in 1/9MR+Gentamicin and a drop of Benzocaine (0.05% in 1/9x MR). Confocal imaging was performed using the Leica DMi8 SP8 microscope with a 40x or 63x oil immersion objective (1.3 NA). Images were captured at 1x, 3x, or 5x zoom and adjusted (brightness and contrast), analyzed, cropped in Fiji, and assembled in Adobe Illustrator software.

All the experiments were repeated three times. All measurements and analyses were performed on at least three embryos per trial, for a total of 3 trials. Sample size, indicated by “n” values, and number of trials, indicated by “N” values, is included in the figure legends. The Fiji freehand selection tool was used to measure ciliary intensity, in which embryos were first thresholded, and the mean gray value within a standard 100×100 pixel box over individual ciliary bundles was then measured and compiled. For analysis of the apical area, the rectangle tool was used to outline the perimeter of five MCCs per embryo to measure the area in microns^²^ and compile the data in Microsoft Excel. Statistical analysis was performed using Prism version 10, where a Welch’s t-test was performed with a significance level of 0.05.

### Flow Analysis in *Xenopus tropicalis* and DAW1 Rescue

To measure mucociliary flow on (uninjected controls and injected) embryos were raised to Stage 28 and anesthetized with benzocaine, 1µL of latex beads was placed at the anterior end of the embryo and visualized under a dissecting scope. If the beads were moved (classified as ‘Flow’) or not (classified as ‘No Flow’) was recorded.

### Cardiac Looping in *Xenopus tropicalis*

The injected *X. tropicalis* embryos that were examined for the presence of mucociliary flow were then allowed to develop to Stage 48 for examination of cardiac formation. The embryos were treated with benzocaine, examined ventrally, and scored for cardiac looping using a light dissection microscope as previously described(Arrigo *et al*., 2025; Sochaka *et al*, 2025). Loop direction is defined by the position of the outflow tract relative to the inflow of the heart: outflow to the right: D loop; outflow to the left: L loop; outflow midline, fails to loop: A loop.

### RNA *in situ* hybridization

*X. tropicalis* embryos (control and MO injected) were collected at Stage 28 for *in situ* hybridization. A digoxigenin-labeled antisense probe for *pitx2* was *in vitro* transcribed with T7 High Yield RNA Synthesis Kit. Embryos were collected and fixed in MEMFA for 2 hours at room temperature and dehydrated for 4-6 hours in 100% EtOH. Briefly summarized, whole mount *in situ* hybridization of digoxigenin-labeled antisense probes was performed overnight, the labeled embryos were then washed, incubated with anti-digoxigenin-AP Fab fragments, and signal was detected using BM-purple, as previously described(Kulkarni & Khokha, 2018).

## Data Availability

All data produced in the present study are available upon reasonable request to the authors

---

Resource list

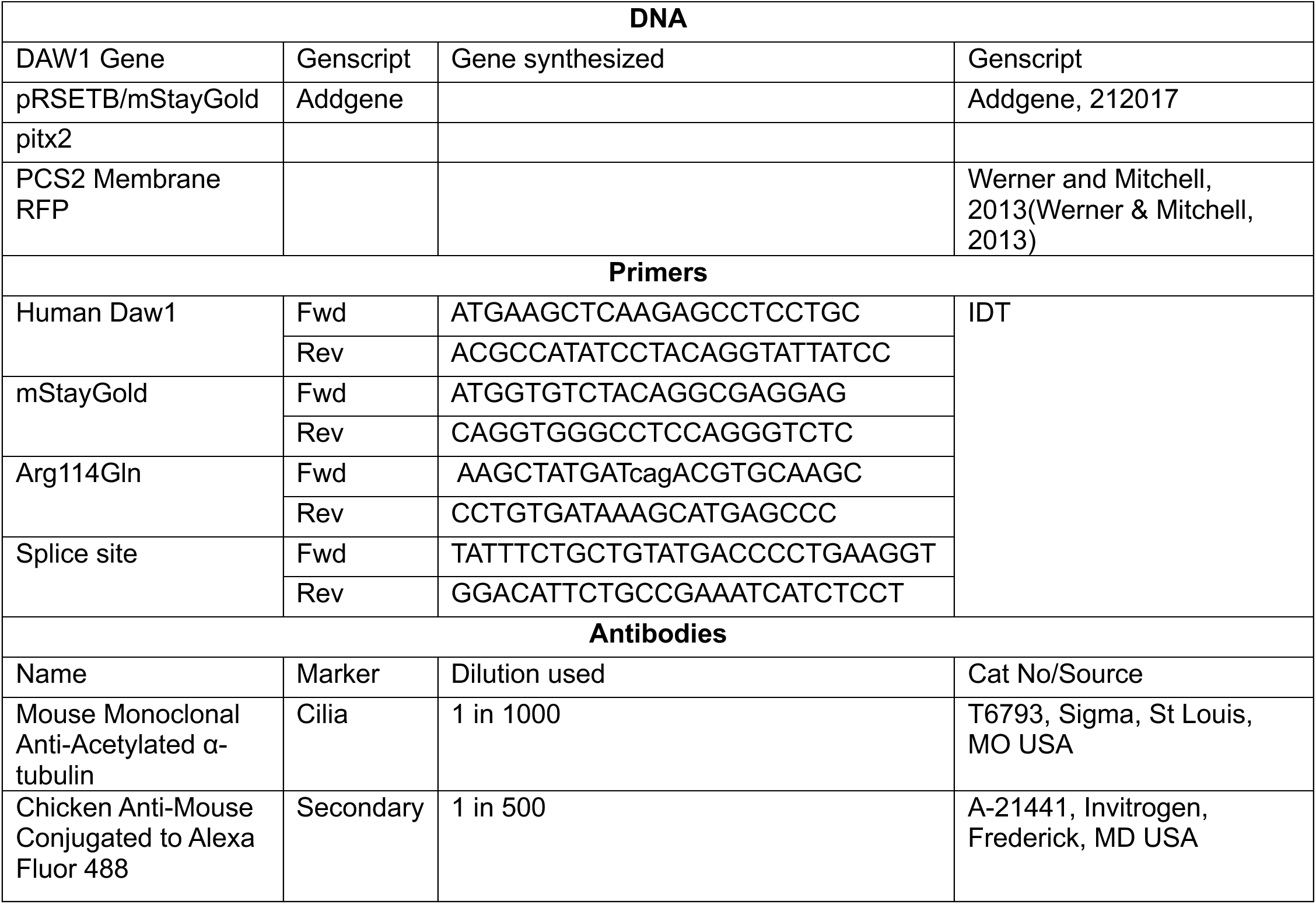
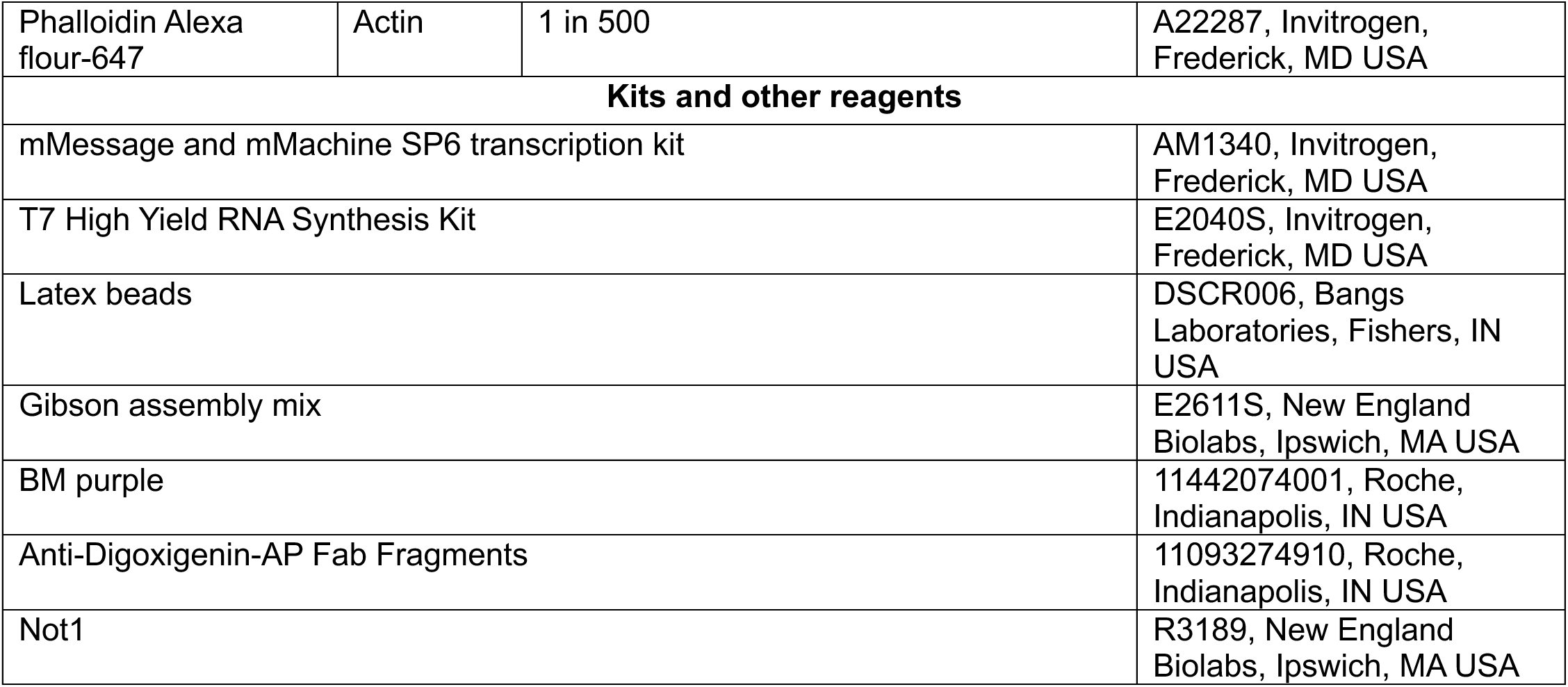

## Supplementary Methods – In silico prediction and structural modeling

The pathogenicity of the DAW1 variants was assessed using multiple computational algorithms accessed through the dbNSFP v4.3 database and its related resources. The following prediction tools were used: Revel, AlphaMissense, MutationAssessor, SIFT, MutationTaster, FATHMM, DANN, MetaLR, and PrimateAI. For the splice-site variant, splicing algorithms SpliceAI, dbscSNV Ada, and dbscSNV RF were applied. All scores were reported in their original scales and interpreted according to tool-specific guidelines. The MetaLR logistic regression ensemble score integrates ten predictors (SIFT, PolyPhen-2 HDIV, PolyPhen-2 HVAR, GERP++, MutationTaster, MutationAssessor, FATHMM, LRT, SiPhy, PhyloP) and 1000 Genomes allele frequencies.

**Table S1:** Summary of all reported DAW1 variants and their associated phenotypes. Previously reported families in Leslie et al., 2022 and Jin et al., 2017 include individuals with homozygous and compound heterozygous variants, presenting with variable features of laterality defects, congenital heart disease (CHD), and respiratory issues. Functional assays in zebrafish distinguished complete loss-of-function alleles (e.g., p.(Asn143Asp), p.(Ser364Thr)) from hypomorphs (e.g., p.(Trp372Cys)). The proband in this study carried compound heterozygous variants (p.(Arg114Gln)/splice-site c.648+1G>A) and exhibited HTX and complex CHD without respiratory disease. *Xenopus tropicalis* assays showed that p.(Arg114Gln) functions as a context-specific hypomorph affecting LRO cilia but not MCCs, while the splice-site variant indicates a complete loss-of-function. These results collectively expand the DAW1-related disease spectrum.

| Family | Zygosity | cDNA Change | Protein Change | Phenotype | Functional Evidence | Reference |
| --- | --- | --- | --- | --- | --- | --- |
| 1 | Hom | c.427A>G | p.(Asn143Asp) | Situs inversus (2 patients, no respiratory); 1 patient respiratory only | Zebrafish LOF (no rescue of cardiac looping or KV motility) | Leslie et al., 2022 |
| 2 | Hom | c.356G>A | p.(Trp119*) | Complex CHD, situs ambiguous, transverse liver, right-sided spleen; no respiratory | Predicted LOF nonsense allele | Leslie et al., 2022 |
| 3 | Hom | c.1091G>C | p.(Ser364Thr) | Complex CHD resembling HTX; respiratory unknown | Zebrafish LOF (no rescue of cardiac looping or KV motility) | Leslie et al., 2022, Jin et al., 2017 |
| 4 | Cmp Het | c.197T>A/c.1116G>T | p.(Leu66*)/p.(Trp372Cys) | Complex CHD resembling HTX; respiratory unknown | Zebrafish partial hypomorph (rescue incomplete) with Trp372Cys | Leslie et al., 2022, Jin et al., 2017 |
| **This study** | Cmp Het | c.341G>A / c.648+1G>A | p.(Arg114Gln) / splice | HTX + complex CHD; no PCD/respiratory disease | *Xenopus*: context-specific hypomorph (R114Q LRO defect, MCC intact); splice LOF | This study |

## Acknowledgement

We thank Dr. Karen Hirschi for providing access to the confocal microscope.

## Author Contributions

DU: Investigation, Analysis, and Manuscript original draft writing and revisions.

AJ: Investigation

SB: Investigation, Manuscript original draft writing

VR: Investigation, Analysis, Manuscript revisions

SCW: Cardiology and imaging

MJT: Patient recruitment and genetic analysis, Manuscript revisions

AG: Clinical insights, Manuscript revisions

CP: Patient recruitment and genetic analysis, Manuscript revisions

AR: Methodology development, Bioinformatic analysis of whole genome data, Manuscript revisions

SSK: Conceptualization, Methodology development, Investigation, Visualization, Supervision, and Manuscript writing and revisions.

## Ethical Approval

Informed consent was obtained from the patient’s family, who were evaluated.

